# Current therapeutic trends for tinnitus cure and control – a scoping review

**DOI:** 10.1101/2021.06.29.21259450

**Authors:** Vatsal A Chhaya, Divya G Patel, Shamik P Mehta, Jignesh P Rajvir, Vinodkumar J Jhinjhuwadia, Pranshuta Sehgal, Kapil M Khambholja

## Abstract

**Introduction:** Chronic tinnitus treatment has been an enigma due to a lack of practice insights. To date, there has been limited evidence-based research on tinnitus therapies available. Our objective for this scoping was to assess the latest knowledge update in the field of tinnitus treatment and/or management and inform the clinical fraternity with evidence-based knowledge.

**Materials and Methods:** We retrieved randomized and non-randomized controlled trials, systematic reviews, meta-analyses, and observational studies from 2014 to 2021(starting from 1 Apr 2021) on chronic tinnitus patients within the context of evidence-based guidelines available on tinnitus therapies. We considered free, full-text, English language articles from PubMed, Cochrane, and Google using an AI-powered evidence synthesis tool named VOODY. We excluded studies on epidemiology, technical objectives assessing other than efficacy/safety outcomes, or review articles. Charted data from selected sources of evidence (related to study characteristics, intervention, sample size, efficacy and safety data, and quality of life-related outcomes as applicable) were presented in the form of graphs, tables, and other visual representations.

**Results:** We retrieved two evidence-based clinical practice guidelines (US and EU regions). Of total 3923 results, 119 underwent screening based on eligibility criteria and 52 were included for the final charting. Out of these 52 articles, three major treatment trends emerged: 1. Acoustic therapies 2. Stimulation Therapies and 3. Herbal, Complementary and Alternative Medicine and Nutrition Therapies. However, none of these were recommended to practice as per the latest EU guideline in 2019 due to no to limited strength of evidence. Other therapies included drugs, cognitive/habituation therapies, and digital interventions.

**Conclusion:** Although tinnitus guidelines did not recommend stimulation therapies, tinnitus research is seen focusing on stimulation. Therefore, it is highly recommended to consider existing clinical practice guidelines and orient tinnitus research focus on digital interventions and multidisciplinary therapies over the standalone therapies for better outcomes.

## Introduction

Tinnitus is an unsolved mystery in the field of otorhinolaryngology because of the unclear mechanisms. It is a clinical condition where patients perceive a ringing sound in their ears, impacting their quality of life. This *“auditory phantom phenomenon”*(1) is often characterized in the form of intermittent or persistent perception of sounds like hissing, chirping, roaring, or whistling. Based on its nature, major types of tinnitus are 1. Subjective 2. Objective 3. Somatic and 4. Neurological.(2) However, regardless of its clinical type, tinnitus can interfere in routine activities of patients and may deteriorate the quality of life. This can take form of anxiety, depression, irritability, sleep cycle disturbances, poor concentration, pain, and even suicidal ideations in extreme cases in tinnitus patients. Tinnitus may not only manifest as a symptom of underlying neurotological, infectious, or cardiovascular diseases but also could be the side-effect of oral drugs with a potential of ototoxicity. Although it is believed that it is caused mostly due to age-related hearing loss and loud noise exposure, in most of the cases, physical cause or mechanism for tinnitus occurrence remains unidentifiable. Due to this challenge associated with disease diagnosis, treatment pathway for tinnitus patients is also blurred with no evidence-based clinical practice. Although Tunkel DE et al.(3) and RFF Cima et al.(4) presented clinical practice guideline (CPG) for tinnitus management, it provided positive recommendation for limited interventions (cognitive behavioral therapy and sound therapy), implying the need of further research on other experimental therapies. These included antidepressants, anticonvulsants, anxiolytics, *Ginkgo biloba*, melatonin, zinc, or other dietary supplements, transcranial magnetic stimulation, and acupuncture. Following the US CPG in 2014, several systematic reviews(5–9) had been published on the Cochrane database with patients suffering from tinnitus as target population, but none of them provided overarching insights on existing therapeutic options for tinnitus due to poor quality or unavailability of data from randomized controlled trials. A typical variant of SLR *aka* Scoping review is a novel and emerging method to inform research priorities to the stakeholders(10). It is a variant of a systematic review with a broader orientation unlike SLR focusing on a particular research problem. Although several scoping reviews(11–24) were conducted by several otorhinolaryngology researchers between 2014-2021, none of them provided holistic insights, indicating a scope of a comprehensive study. This motivated us to explore the latest evidence and to assess the scope of knowledge update with a broader approach for tinnitus treatment and/or management. Another purpose behind this scoping review was to provide an overarching overview of experimental tinnitus therapies to the clinical fraternity within the context of evidence-based CPGs available in public domain.

We developed our ScR to address following research question(s).

1. ***Does any updated evidence exist for treatment or management of Tinnitus? If yes, what are the key therapies studied in the same?***
2. ***Does current tinnitus research align with research and practice recommendations in existing clinical practice guidelines?***
3. ***What could be the future research trends within the context of existing CPGs for tinnitus treatment and/or management?***

## Material and Methods

We followed Joanna Briggs Institute (JBI) methodological guidance for Scoping Reviews(25) to prepare our report.

### Protocol registration

The initial version of the protocol for this study was registered at the open science framework registry (OSF), registration *DOI: 10*.*17605/OSF*.*IO/R8D39*. Based on several inputs from the global subject matter experts, we modified it to better comply with latest reporting guidance(26).

### Eligibility Criteria and Information Sources

Peer-reviewed journal records(studies) matching with all of the following conditions were included: 1. Published in the *English* (due to no translator(s) available) on *humans* between 2014-2021. 2. Free, full-text available (due to no funding available). We considered Joanna Briggs Institute (JBI) Scoping Reviews mnemonic framework – *Population, Concept, Context (PCC) -* for the eligibility assessment of retrieved records (Figure 1). Accordingly, studies containing any information on efficacy and/or safety outcomes for therapy(ies) in tinnitus patients were deemed eligible for inclusion. We excluded studies focusing on diagnosis aspects, specific risk groups, epidemiology, or any technical objective focusing other than efficacy outcome(s). Eligible study designs for our ScR included systematic reviews, meta-analysis, randomized controlled trials (RCTs), and observational studies. We disregarded review articles based on reliability issues. The rationale for selecting timeframe between 2014-2021(*Search Initiated on 1 Apr 2021)* for our ScR was to identify the tinnitus research occurred after the release of clinical practice guideline (CPG) by Tunkel et al.(2014)(3).

**Figure 1.**
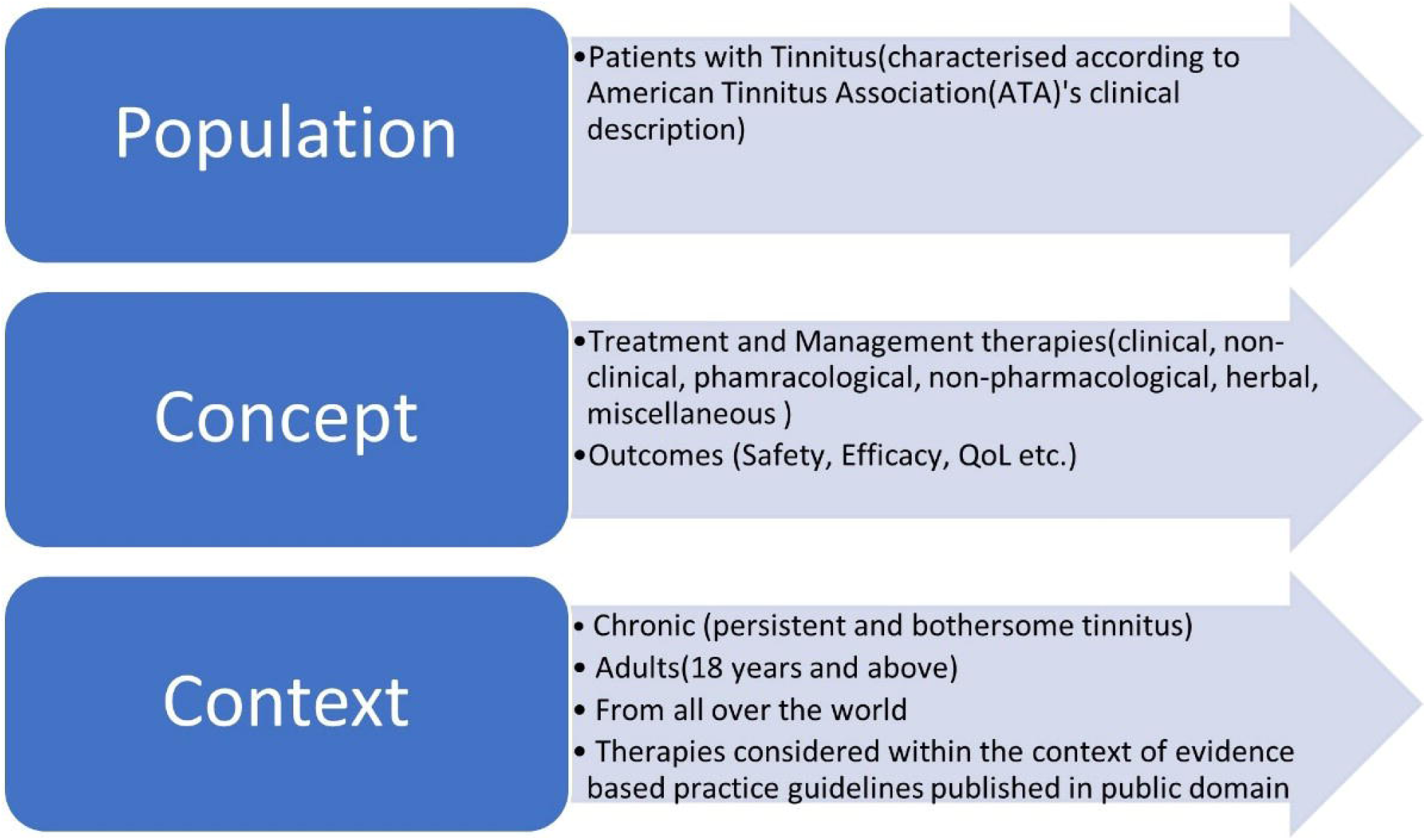
Scoping Review Mnemonic (PCC) Framework

### Search Strategy

VC and DP identified relevant key terms and developed detailed search strategy from a preliminary search. The final search strategy included two clinical literature databases (PubMed and Cochrane Library) and was comprised of both medical subject headings (MeSH) terms and/or text words related to tinnitus and various interventions studied for tinnitus treatment. Detailed search strategy for PubMed is provided in Table 1.

**Table 1 :** Sample search strategy for PubMed

| Search Strings | Search Filters |
| --- | --- |
| ((((((((((((((((tinnitus[MeSH Terms]) ) OR (clicking tinnitus[MeSH Terms])) OR (induced tinnitus, noise[MeSH Terms])) OR (leudet s tinnitus[MeSH Terms])) OR (leudet tinnitus[MeSH Terms])) OR (leudet's tinnitus[MeSH Terms])) OR (disease, meniere's[MeSH Terms])) OR (hyperacusis[MeSH Terms])) OR (hearing loss[Text Word])) OR (subjective tinnitus[Text Word])) OR (objective tinnitus[Text Word])) OR (neurological tinnitus[Text Word])) OR (somatic tinnitus[Text Word])) OR (drug induced tinnitus[Text Word])) OR (surgery induced tinnitus[Text Word])) AND (((((((((((((((((((therap*) OR (management)) OR (treatment)) OR (care)) OR (surgical)) OR (non-surgical)) OR (medical)) OR (pharmacological)) OR (non-pharmacological)) OR (herbal))) OR (auditory stimulation[MeSH Terms])) OR (magnetic stimulation, transcranial[MeSH Terms])) OR (vagus nerve stimulation[MeSH Terms])) OR (stimulation, deep brain[MeSH Terms])) OR (direct current stimulation[Text Word])) OR (surger*)) OR (outcome*)) OR (severity)) OR (scale)) OR (drug*)) OR (quality of life)) OR (HR-QoL)) OR (QoL)) OR (minimum masking level)) OR (MML)) | Clinical Trial,<br>Meta-Analysis,<br>Observational<br>Study,<br>Randomized<br>Controlled<br>Trial, Review,<br>Systematic<br>Review,<br>Humans, from<br>2014 - 2021 |

The PubMed-based records retrieval was performed using an AI-powered tool named VOODY (*Genpro Research Inc*.) (DP and VC). This was supplemented by a targeted search was performed from additional sources like Google, American Tinnitus Association, British Tinnitus Association, All Indian Institute of Speech and Hearing (AIISH), and Joanna Briggs Institute Evidence Synthesis to retrieve if any relevant information available. Also, an exclusive targeted search for tinnitus-related scoping review(s) was performed using the VOODY.

### Selection of source(s) of evidence

Two independent reviewers (DP and PS) performed the screening and selection of retrieved records. Any discrepancy regarding screening and/or inclusion was resolved through consultation with the third reviewer (KK).

### Data charting

The data charting instrument/template was prepared by referring to JBI Scoping Review Data Extraction Temple and was piloted on random five articles (VC). Following necessary modifications post-pilot work, DP and PS performed the charting using a Microsoft Excel spreadsheet. SM, JR, and KK cross-verified the charted data with the help of source records. Data discrepancies were resolved by mutual discussion and/or further adjudication from the third reviewer (VJ) if needed. The abstracted data items included study type, sample size, patient characteristics(age, gender, region, tinnitus type), interventions studied to treat tinnitus patients(pharmacological, surgical, various stimulations, herbal or any other treatment options), study objective(primary and secondary if given), study outcome(s)(effect on tinnitus perception, severity, loudness, hearing loss, quality of life, adverse events), and conclusion remarks/recommendations from the study author(s) as applicable. Charted, verified data was then presented in form of tables and graphical visualizations where possible.

## Results

Total 3923 records were retrieved based on the final search strategy. After removal of duplicates (n = 18) and exclusion based on pre-screening search filters (study type, time frame, species and full-text availability) (n = 3094), 811 records underwent first level of screening based on title/abstract and/or relevance with the study topic. Of these, 119 records qualified for eligibility screening. Based on various reasons (different study design, mismatch with research objective, different study population, or any other reason proving ineligibility of respective record), 67 records were excluded, yielding 52 records for inclusion in our review. Our targeted search with relevant key words (Tinnitus therapy guidelines, Tinnitus management white paper, Tinnitus cure, Evidence based consensus for tinnitus) revealed zero free-text articles from ASHA wire, AIISH websites due to paid access. However, two evidence-based clinical practice guidelines and a scoping review (retrieved via Google) were included in final review. A detailed description of the study selection process is shown in PRISMA-ScR flow diagram (Figure 2).

**Figure 2.**
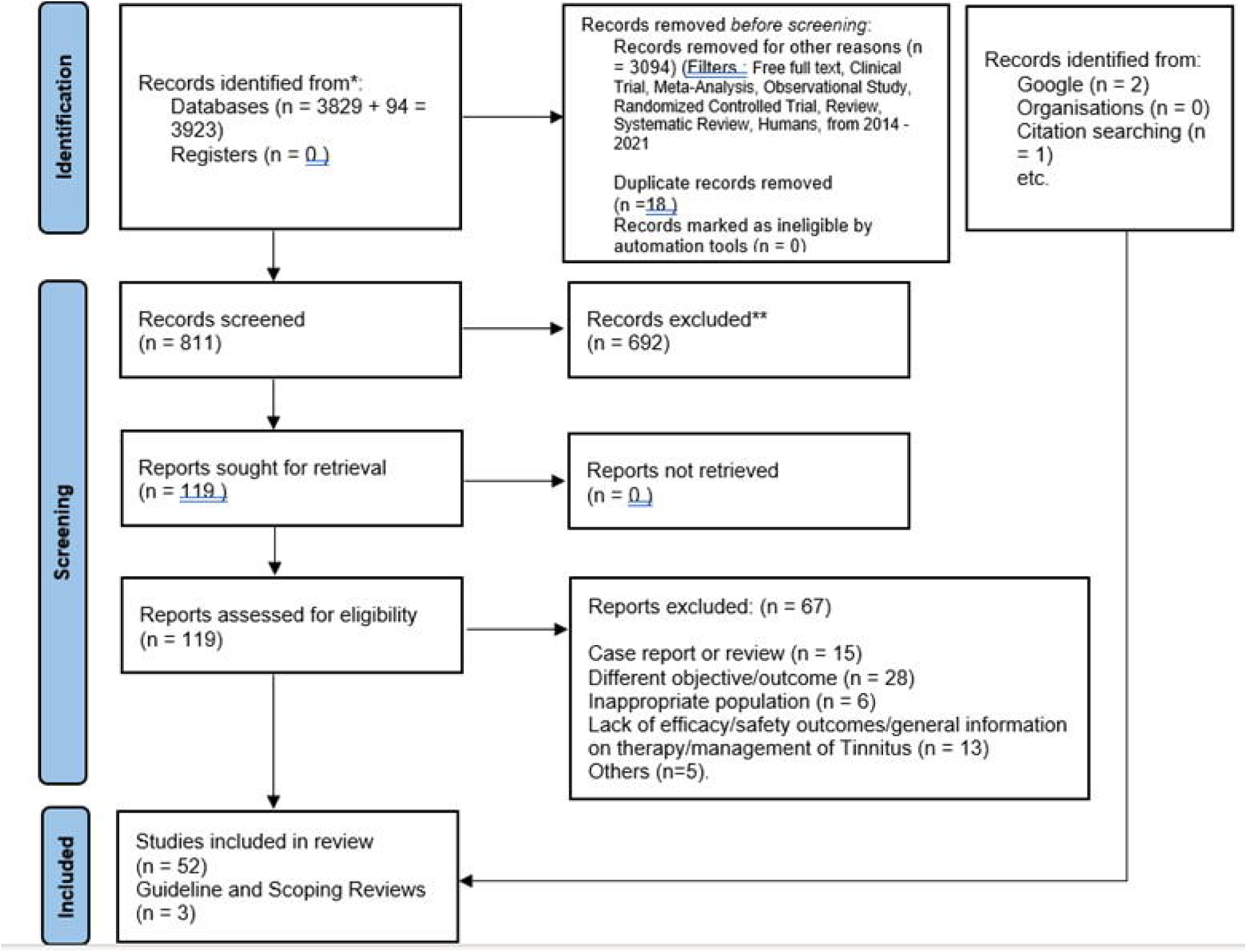
PRISMA-ScR flow diagram

Fifty-two total included records comprised of 16 SLR/MAs (5–9,27–37)(30.18%), 34 RCTs (64.15%)(38–71), one non-randomized CT(72), and one observational study(73) (5.66%). (Figure 3). Three major treatment options from collected evidence included: 1. Electric/magnetic stimulation (28–32,34,36,41,47,50,52–54,57,61,64,65,67,68) 2. Herbal, Complementary & Alternative Medicine (CAM), and Nutritional therapy (8,19,37,39,40,42,49,59,60,63), and 3. Acoustic therapy (related to sound) (7,9,38,51,55,62,71–73). Other interventions included behavioural, digital, and drug or medical gas related type (Figure 4). Notably, half of the total number of SLRs (50%, n = 8/16) also corresponded to stimulation-based interventions. Two authors (DP and PS) performed data charting from the selected source of evidence focusing on following characteristics: participants, sample size, interventions, study origin, type of study, tinnitus measurement tool, and efficacy, safety and/or QoL outcomes.

**Figure 3.**
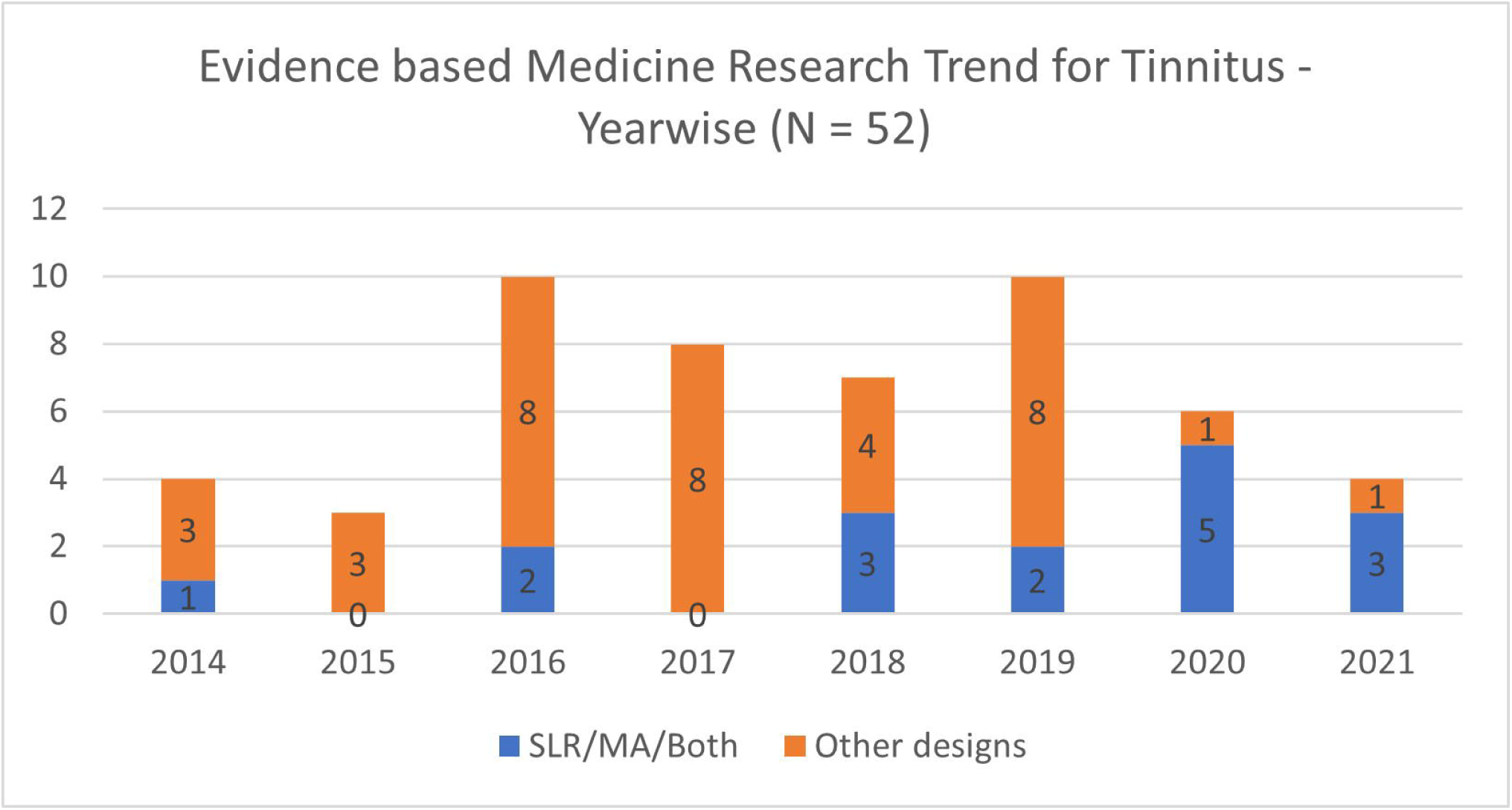
Evidence-based tinnitus research trend between 2014-2021

**Figure 4.**
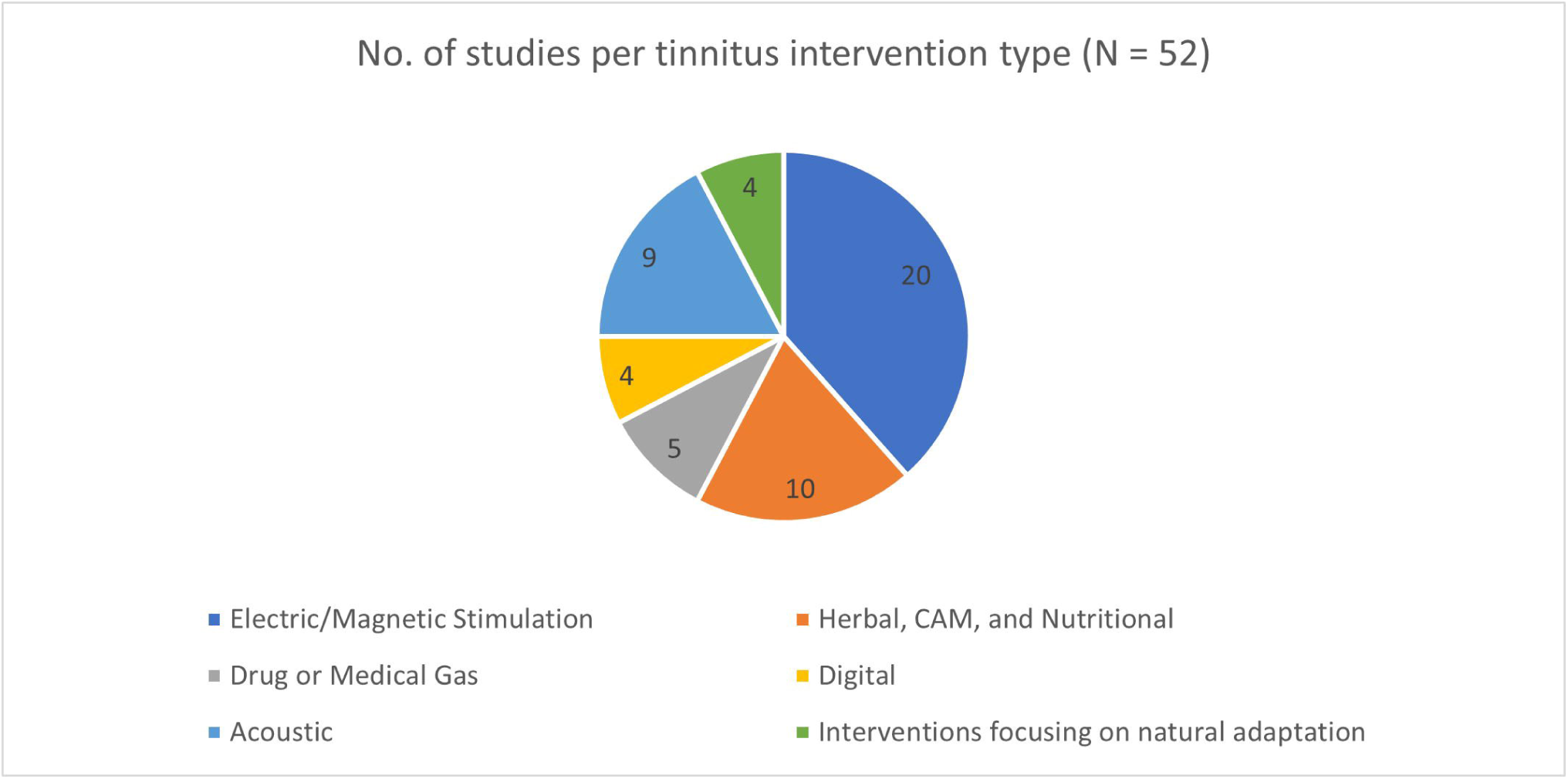
Study distribution based on type of tinnitus interventions

### Key Findings

#### Population

12 and 40 studies specified patients with hearing loss plus tinnitus and tinnitus only, respectively for their inclusion. The sample size for RCTs ranged from 12 patients to 374 patients. 20.58% of total RCTs (N = 7) mentioned sample size more than or equal to 100 patients. Similarly, sample size for SLR evidence widely ranged from 74 patients to 4165 patients. Majority of studies in present ScR were from developed countries(Germany, USA and UK) (N = 26).

#### Concept

Of total 24 experimental therapies reviewed in our ScR; no additional therapies were found to those considered in existing CPGs from the US and Europe.

#### Acoustic Therapies

It included notched music therapy, customized sound therapy, sound generators, acoustic coordinated reset neuromodulation. Notched music therapy studies presented equivocal evidence with both significant and non-significant results. Similarly, sound generator RCTs demonstrated significant effect; however, SLRs on sound generator (2018) and hearing aids (2014) did not provide any supporting, strong evidence except for small clinical significance from either. An observational study on acoustic coordinated reset neuromodulation also demonstrated clinically and statistically significant changes in TBF-12 and CGI-I7 scores.

#### Stimulation Therapies

It included cochlear implant, vagus nerve stimulation, repetitive transcranial magnetic stimulation and its parameters, non-invasive brain stimulation, transcranial random noise stimulation, and transcranial direct current stimulation. Only eight out of 18 studies (%) that measured effect on tinnitus severity demonstrated significant effect of respective stimulation techniques. Remaining studies focused on operational aspects and not efficacy outcomes.

#### Herbal, CAM and Nutritional Therapies

It included Gingko Biloba extract EGb 761, antioxidants, acupuncture, zinc supplementations, and Vitamin B12. Three (including SLR also) out of four studies that studied acupuncture therapy showed no beneficial effect. However, Gingko Biloba studies showed significant reduction on respective tinnitus measurement scales after 3 months. Of three nutritional therapeutic option studied, only Vitamin B12 study reported significant results in patients with vitamin B12 deficiency.

The detailed characteristics charted from individual source of evidence is depicted in table 2 (Supplement file 1).

**Table 2 :** Characteristics for individual sources of evidence (data charting) Attached as a separate MS-Excel spreadsheet..

The comparative analysis between recommendations from European and US clinical practice guidelines (CPGs) did not result in any noticeable finding as shown in table 3.

**Table 3 :**
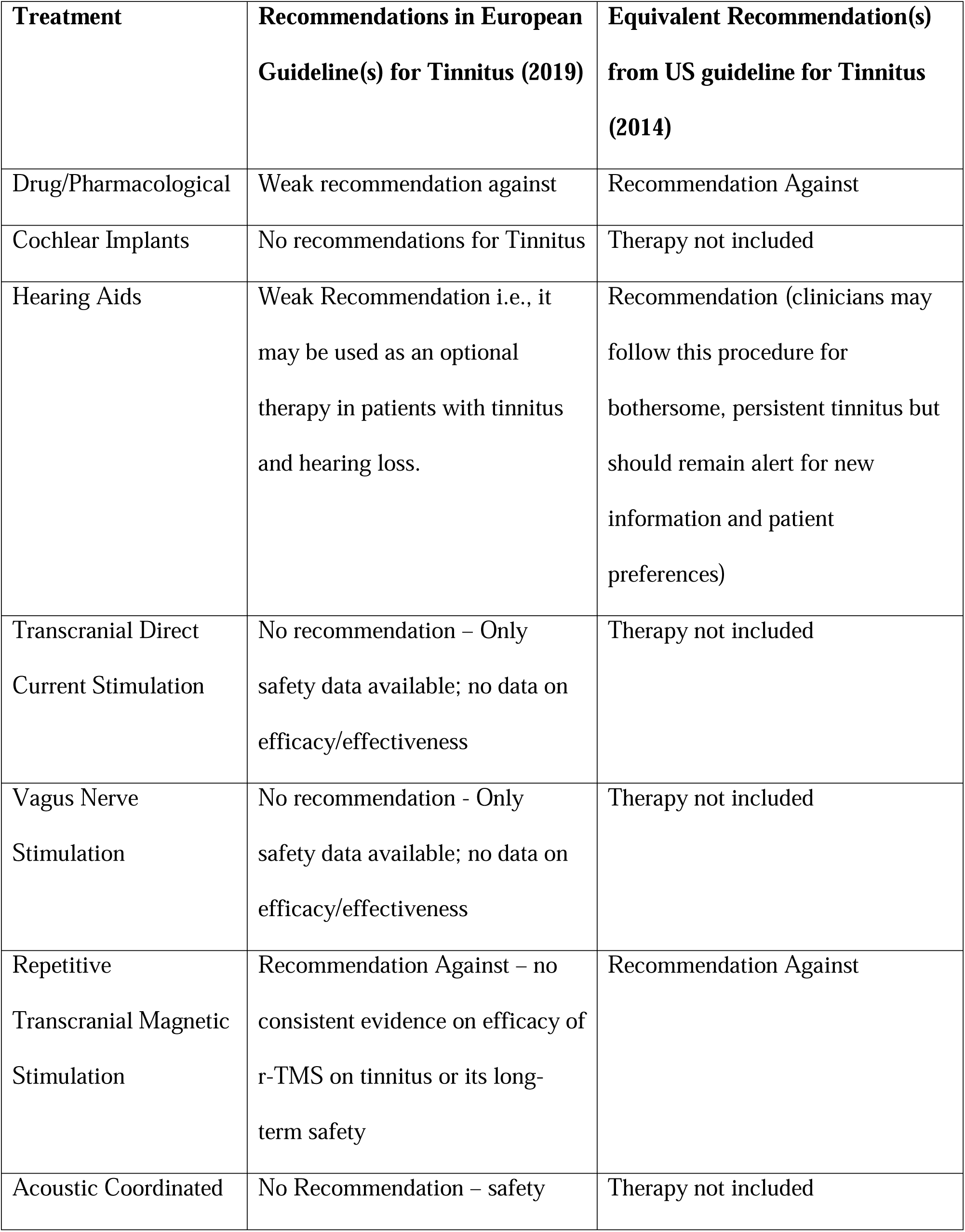
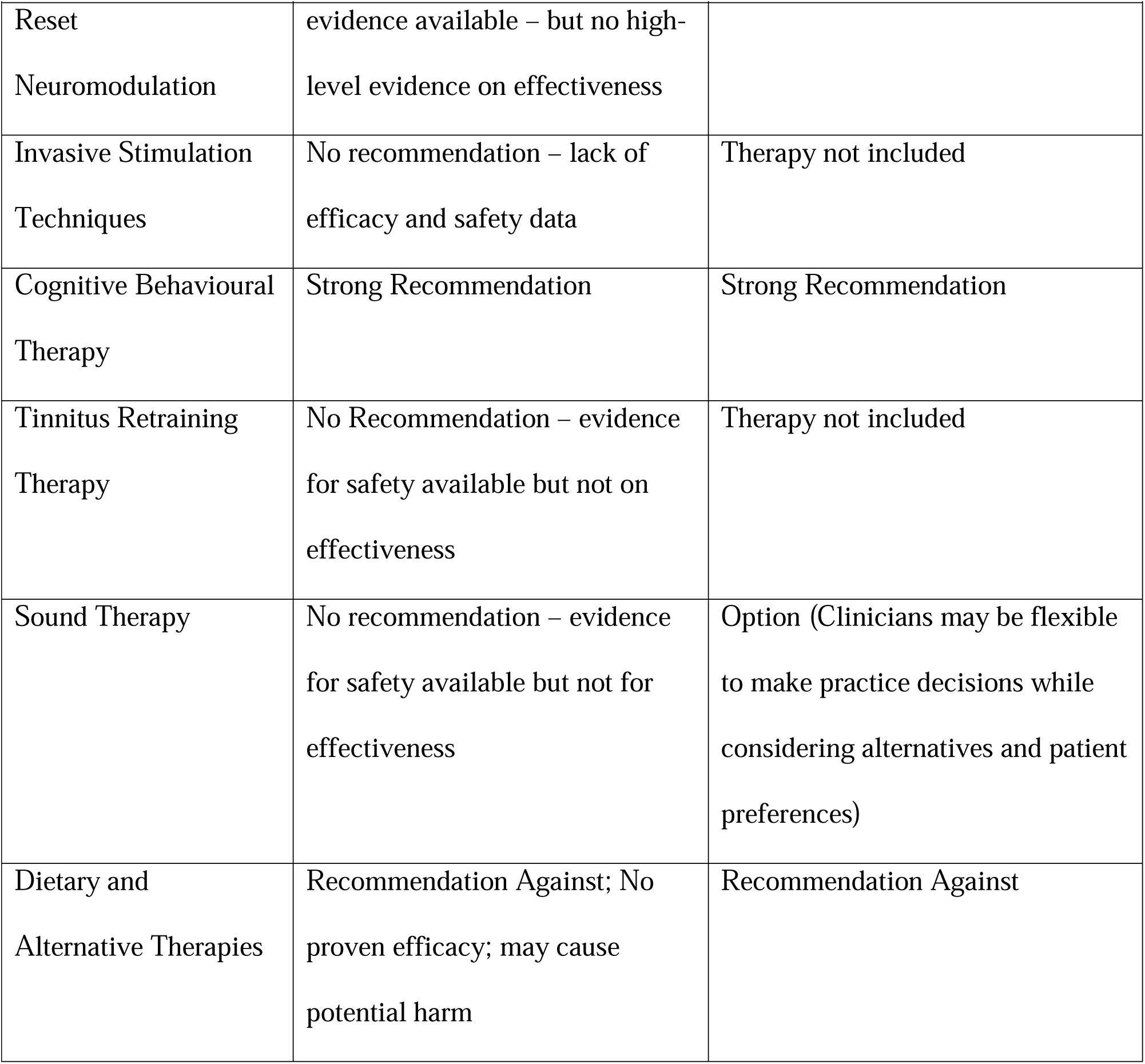
Comparison between practice recommendations found in US and EU CPGs regarding various tinnitus therapies

## Discussion

We performed a scoping review to broadly explore the treatment landscape for chronic tinnitus especially after the release of 2014 evidence-based tinnitus management guideline. Scoping review is an emerging research method to assess the evidence gaps and to obtain a bird eye view on any specific topic. Although several scoping reviews are available on tinnitus, they either focused on individual therapy or had different objective/population. Our preliminary literature search identified two guidelines from European and USA regions(3,4). However, the only scoping review exploring tinnitus therapy a whole did not include any of them. Also, Makar S et al. 2017(18) did not refer to any standard reporting checklists to ascertain the quality of reporting. Therefore, Our ScR is first in its kind with well-defined research objective (to explore tinnitus research landscape within the context of evidence-based guidelines in 2014 & 2019) and in concordance with PRISMA-ScR checklist 2020. Aligning with our research objective, we kept timeframe for our literature search from 2014-2021 starting from 1 Apr 2021. Being a scoping review with broad concepts regarding study designs, we included SLR and SLR-MA, RCTs, non-randomized CTs and observational studies. However, we did not consider case report and review articles due to their low scientific hierarchy. This being a non-funded project, we limited our search to free-full-text articles only. A scoping review by Makar et al. (18) described the interventions with intended cognitive effect on tinnitus, including counseling, tinnitus masking, TRT, CBT, relaxation, and attention diversion in tinnitus patients. Complementing the same, our study described all experimental therapies within the context of evidence-based guidelines for tinnitus from 2014 and 2019(3).

We included 55 studies (including two evidence-based guidelines for treatment and/or management of tinnitus and a scoping review). Our ScR did not include any additional therapy than those mentioned in European guidelines, indicating the dearth of novel therapies for tinnitus in recent past. Surprisingly, there were limited studies from Asian region with no study from India except for one ScR. This demonstrated limited awareness and/or interest towards tinnitus research in low and middle-income countries like India. Collectively, none of therapies except for cognitive behavioural therapy was found with significant findings with strong recommendations, demonstrating a scope of evaluating methodological inconsistencies for the interventions other than CBT.

Compared with guideline by Tunkel 2014, there were six new therapy modalities found in European guidelines, reflecting the evolution in tinnitus research over the period of time. Contradictorily, we found majority studies focusing on interventions related to stimulation therapies and acoustic therapies despite no strong recommendations for such therapies in both evidence-based guidelines, pointing towards plateaued phase of tinnitus research.

Out of 12 therapy modalities considered in the European guideline, seven presented no recommendation owing to the lack of safety/efficacy/both data. This shows unmet research need and warrants well-designed studies for reforming current practice models.

Apart from evaluation of efficacy and safety of an intervention for tinnitus as a whole, we also found multitude of research objectives for same therapy. For example, Dong C and team evaluated low frequency rTMS whereas Schoisswohl S et al. and Sahlsten H et al. studied effect of change in technical parameters in rTMS and effect of navigation in RMS efficacy, respectively. Unlike systematic review, scoping review aims to broadly capture relevant information. Therefore, such bundle of studies with multiple research objective(s) for the same intervention was possible to add in our ScR.

However, we could not extract any actionable insights out of available studies assessing pharmacological, CAM or similar interventions for tinnitus. The possible reason as experienced by many tinnitus researchers could be the inherent disease complexity. Given the increasing use of artificial intelligence in proteomics and metabolomics for lead identification, we encourage tinnitus researchers to decipher complex tinnitus mechanisms and to explore new drugs with AI-induced target identification or AI-induced frequency modulation. Another important thing that we noticed was a huge evidence gap in terms of reporting secondary outcomes i.e., depression, anxiety and QoL. Only ten studies reported improvement/no improvement in QoL, rendering dearth on similar aspects for the rest of the interventions.

Lastly, although none of the guidelines provide in-depth information on use of digital means to deliver interventions for treating or managing the tinnitus, our ScR analyzed four studies using internet-based or mobile applications-based solutions for delivering CBT with promising outcomes, informing the future cost-effective tinnitus research especially post COVID 19 pandemic.

One of the major limitations that we acknowlege is the modification in the previous version of registered protocol to comply with standard methodological guidance for ScRs and to accommodate inputs from experts following pre-print upload on MedRxiv and OSF registration. (*Summary of Changes will be available upon request)*. Secondly, we found no study focusing on multidisciplinary therapy approach for tinnitus treatment and/or management despite its benefit and proven results in the patients with tinnitus.Also, we could not assess a few technical nitty-gritties associated with several therapies like repetitive transcranial magnetic stimulation, multi-frequency matching and therapies involving frequency fundamentals because of absence of acoustic technologist in our research team.Lastly, lack of translation facility did not allow us to capture non-English literature, which may be considered as a future scope of improvement.

## Conclusion

Our scoping review is first in its kind in the domain of tinnitus research. It provided holistic insights about available therapies for tinnitus treatment and management since the release of evidence-based guidelines in the US and thereafter. Considering the evidence gap from low and middle-income countries, otorhinolaryngology professionals and acoustics professionals from countries like India are encouraged to undertake large scale burden of disease studies and interventional research on various therapies recommended in global CPGs. Given the lack of high-quality efficacy and safety data, it would be worthwhile to promote primary research with meticulous methods onto acoustic technologies and stimulation therapies especially those of lesser harm. Implementation research focusing on effectiveness of multidisciplinary approach is highly recommended over the assessment of therapeutic potential of individual therapies. Digital interventions for CBT delivery may provide newer insights in tinnitus treatment and management, however, requiring robust evidence for health system integration.

## Data Availability

All the data charted in the study is available in public domain and can be accessed using bibliography.

